# COVID-19 related social distancing measures and reduction in city mobility

**DOI:** 10.1101/2020.03.30.20048090

**Authors:** Amyn A. Malik, Chandra Couzens, Saad B. Omer

## Abstract

In the absence of any pharmacological intervention, one approach to slowing the COVID-19 pandemic is reducing the contact rate in the population through social distancing. Governments the world over have instituted different measures to increase social distancing but information on their effectiveness in reducing mobility is lacking. We analyzed the mobility data from 41 cities to look at the effect of these interventions. The median mobility across cities on March 2, 2020 was 100% (IQR: 94%, 107%), which decreased to a median of 10% (IQR: 7%, 17%) on March 26, 2020. We found that the mobility decreased on average by 3.4% (95%CI: 3.3%, 3.6%) per day from March 2 through March 26. Social distancing measures decreased the mobility by an additional 23% (95%CI: 20%, 27%). Our study provides initial evidence for the reduction in mobility in cities instituting social distancing measures.

---

A novel coronavirus disease, COVID-19 is causing a global pandemic with approximately 800,000 cases as of March 30, 2020.^1^ In the absence of any pharmacological intervention, one approach to slowing the pandemic is reducing the contact rate in the population through social distancing.^2^ Governments the world over have instituted different measures to increase social distancing but information on their effectiveness in reducing mobility is lacking. Here we analyze the mobility data from 41 cities to look at the effect of these interventions.

We downloaded mobility data from the Citymapper Mobility Index (CMI) from March 2, 2020 to March 26, 2020 for our analysis.^3^ Citymapper is a public transit and map service spanning 41 urban cities globally. It has over 20 million users and helps optimize routes for public transit, biking, walking and ridesharing applications; it does not support personal automobile navigation. The CMI is based on planned trips on the Citymapper application.^3^ We tabulated the data on implementation of governmental social distancing measures between March 2 and March 26 from official government and media websites. We classified a city to have instituted social distancing measures if non-essential businesses were closed; these measures were further classified as moderate or intense based on the intensity of closure. We estimated the effect of time and social distancing measures using a multilevel mixed-effects linear regression model.

We had a total of 1,025 observations across 41 cities (25 observations per city) in our dataset. The median mobility across cities on March 2, 2020 was 100% (IQR: 94%, 107%), which decreased to a median of 10% (IQR: 7%, 17%) on March 26, 2020 (Figure 1). We found that the mobility decreased on average by 3.4% (95%CI: 3.3%, 3.6%) per day from March 2 through March 26. Social distancing measures decreased the mobility by an additional 23% (95%CI: 20%, 27%). There was no difference in mobility reduction based on whether the measures were moderate or intense.

**Figure 1.**
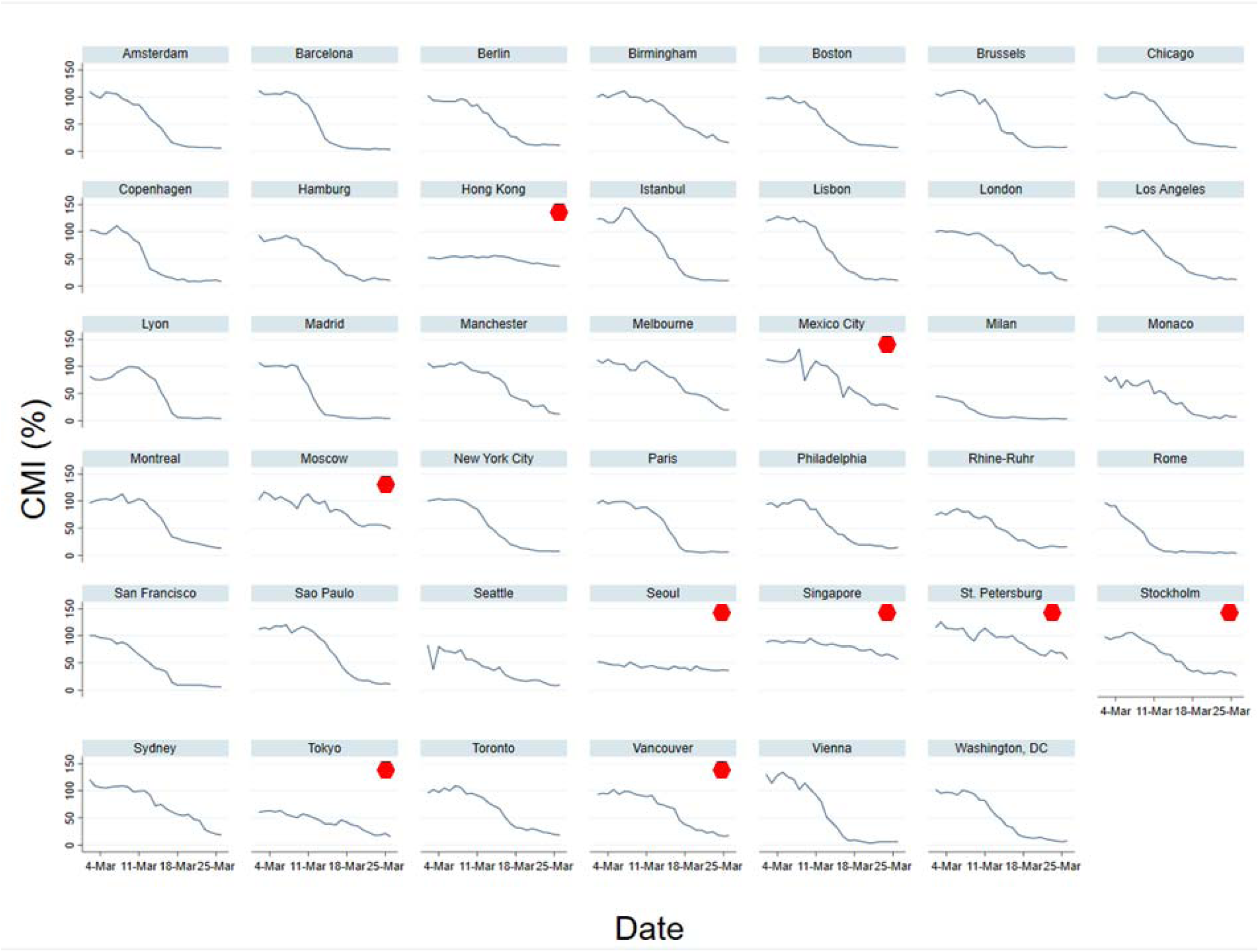
Trend in City Mobility Index (CMI) in 41 cities compared to baseline. 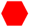 Cities that did not institute social distancing measures between March 2 and March 26, 2020

The reduction of 3.4% per day in mobility can be explained by the repeated emphasis on social distancing by public health authorities. In addition, social distancing measures introduced by governments reduced the mobility in the concerned cities by 23%. Our findings may not be generalizable to the population that does not use public transport. However, since there is a higher risk of transmission in public transport versus private vehicles, our findings may represent decrease in mobility in an epidemiologically relevant group.

## Data Availability

All data is publicly available and can also be requested from the corresponding author.

## References

1. An interactive web-based dashboard to track COVID-19 in real time. Lancet Infect Dis (2020), 10.1016/S1473-3099(20)30120-1

2. Imperial College COVID-19 Response Team. Impact of non-pharmaceutical interventions (NPIs) to reduce COVID19 mortality and healthcare demand. 16 March 2020. London: Imperial College, 2020 (https://www.imperial.ac.uk/media/imperial-college/medicine/sph/ide/gida-fellowships/ImperialCollege-COVID19-NPI-modelling-16-03-2020.pdf)

3. Citymapper Mobility Index. London: 2020 (https://citymapper.com/cmi)

